# Addressing the challenges of estimating the target population in calculation of routine infant immunization coverage in Kenya

**DOI:** 10.1101/2025.01.30.25321415

**Authors:** Christine Karanja-Chege, Ambrose Agweyu, Fred Were, Michael Boele van Hensbroek, William Ogallo

## Abstract

Target population estimation for immunization coverage calculations through extrapolation of annual births from census data is often inaccurate. This study aimed to evaluate the accuracy of the traditional census extrapolation method in comparison with three alternative approaches: the Cohort-Component Population Projections Method (CCPPM), using the Expanded Program on Immunisation (EPI) numerator as a denominator, and estimates derived from first antenatal care clinic (ANC1) visits.

We obtained target population estimates from 1999 - 2023 using all 4 methods with data for ANC1 available only for 2020-2023. We assessed the accuracy of the estimates for 2003 to 2018 by computing the Mean Absolute Error (MAE), Mean Absolute Percentage Error (MAPE) and the Pearson Correlation Coefficient (r), excluding outliers. A sub-analysis for the period 2020-2023 included ANC1 data.

The CCPPM method had the largest population estimates while the census-based method had pronounced discontinuities at the census years. The CCPPM method compared to the DTP1 doses was associated with the greatest error magnitude (MAE = 212917.19 and MAPE = 18.18) while the DTP1 doses and census-based methods showed the smallest error (MAE = 44317.16 and MAPE = 3.77). Sub-analysis of target populations for the period 2020-2023 showed similar upward trends except for the census-based method which exhibited a relatively flat and significantly divergent trajectory. Comparison between the ANC1 and DTP1 doses showed the strongest linear correlation (r = 1.00).

The results reveal significant inaccuracies in the current target population estimation methods which may have serious implications on immunisation coverage assessments. Immunisation programs should utilise diverse sources of data and triangulate results to approximate the true population. Additionally, there is an urgent need to come up with innovative approaches to estimating the target populations for immunisation.

## Introduction

Immunization coverage is defined as the number of people who have received a specific vaccine within a specified target population [1]. In the administrative method of estimating immunisation coverage employed by most EPI programmes, the number of doses of a specified vaccine or vaccines (numerator) is expressed as a proportion of a defined target population (denominator) [2]. The aggregated data is then conveyed to the WHO and UNICEF Estimates of National Immunisation Coverage (WUENIC) platform using the Joint Reporting Form (JRF) every year [3].

Obtaining the record of births for the specified period from the national civil registry is the most accurate method of obtaining the target population for routine infant vaccination.

Unfortunately, in most low resourced countries, including Kenya, the rate of birth registration is low. The Kenya Vital Statistics Report (2021) documented that 14% of Kenyan children born in 2021 were not registered [4].

Target population estimates for intercensal years are therefore obtained by extrapolation of census data using the annual population growth rates [5]. To calculate the coverage for the vaccines given at birth or close to birth such as BCG, annual birth cohorts form the target population while for the rest of the vaccines given in infancy, the estimated number of surviving births under 1 year is used as the target population [6].

Estimating the target population using census projections is highly prone to error particularly at the sub-national level as population estimates do not account for factors like migration and cases where vaccination is obtained from different health facilities. This leads to denominator inaccuracies resulting in coverages of more than 100%. [6].

As immunisation coverage rises over time, a high magnitude of error in the target population estimation results in a greater error in immunisation coverage calculations. Incorrect coverage reporting can give false information regarding the state of national EPI programmes making them fail to allocate resources for interventions to improve coverage [6].

To address the target population estimation challenges, WHO developed a denominator guide in 2015. The guide recommends comparison of 2 or more alternative target population estimation methods with that used by the EPI program in order to enhance accuracy [6].

The National Vaccines and Immunisation Programme in Kenya employs a consultative process involving county EPI managers, Health information systems staff and other key immunization stakeholders including global partners to arrive at a consensus regarding the target population estimates. Census projections data from the Kenya National Bureau of statistics are compared with other sources such as the United Nations Division of Population (UNDP) population estimates.

Since the inception of the Kenya Expanded Program on Immunisation (KEPI) in 1980, national coverage has shown a progressively upward trend, remaining consistently above 80% from 2018 [7]. It is therefore of paramount importance that target population estimates be as close as possible to the true value to avoid inaccurate immunisation coverage reporting that may give misleading information to policy makers. The objective of this paper is to compare the different methods of estimating the target population for routine infant immunisation, demonstrating the need to utilise different sources of population data to improve accuracy.

## Methods

The target population estimation methods that we investigated in this study were: the census-based population projections, the Cohort-Component Population Projections Method (CCPPM) and the Expanded Program on Immunisation (EPI) numerator as denominator and attendance of the first antenatal care clinic (ANC-1) methods. We visualised the target population trends from 1999-2023. Data for the ANC-1 method were only available for the years 2020-2023, and analysis was therefore confined to that period.

### Census-based Population Projections

The Census-based Population Projections method involved estimating annual target populations from the most recent census year, using the annual growth rates available from the Kenya National Bureau of statistics (KNBS) [8]. Using data obtained from the 1999, 2009 and 2019 censuses available on the KNBS website, we extracted the national population of surviving infants 0-1 year. We then computed the national annual target population estimates using the population growth rates provided in the census reports over the 3 census periods and extrapolated to 2023.

### The CCPPM method

The Cohort-Component Method for Projecting Population (CCMPP) employed by UNDP utilises fertility, mortality and migration data disaggregated by age and sex to formulate the population estimates. It is fairly accurate compared to most other population estimation methods [9, 10] and is recommended as a credible alternative population data source.

We extracted the population of annual live births and surviving infants 0-1 year for the period 1999 to 2023 from the UNDP annual World Population Prospects report (2024).

### The EPI Numerator as Denominator Method

The EPI numerator as denominator method uses the number of vaccine doses administered to estimate the target population. Vaccines such as BCG or the first dose of Diphtheria-Tetanus-Pertussis (DTP-1) administered at birth or close to birth have been shown to have a high coverage rate. The WUENIC estimates of BCG and DTP-1 vaccine coverage in Kenya from 2000 to 2023 have been over 90% with DTP-1 coverage sometimes out-performing BCG [11]. It is assumed that the number of BCG and DTP-1 doses administered per year closely mirrors the national birth cohorts and surviving births up to one year respectively [6].

From the EPI data available on the WUENIC portal, we obtained the annual number of BCG and DTP-1 vaccine doses administered in Kenya at national level over the 3 census periods from 1999-2023.

### The first antenatal care visit (ANC-1)

The numbers of women attending the first antenatal care clinic in a pregnancy has been used as a measure of annual expected births. This parameter can be used to estimate the target population for birth vaccines as ANC1 coverage rates were shown to be over 90% in the Kenya Health Sector Strategic and investment plan 2018-2023 [12]. We extracted the ANC-1 coverage data for 2020-2023 available on the UNICEF data portal [13].

### Comparison of paired target population estimation methods

We compared pairs of target population estimation methods by calculating the magnitude of error and the linear correlation.

We measured the magnitude of error by calculating the Mean Absolute Error (MAE) and Mean Absolute Percentage Error (MAPE) associated with paired combinations of each of the methods. We also evaluated the linear correlation between the methods by calculating the Pearson Correlation Coefficient.

We confined the analysis for the census-based population projections, the CCPPM and the EPI numerator as denominator methods to the years 2003 - 2018 due to the presence of outliers in the other years. Since data for ANC1 was only available from 2020-2023, we included this variable for that period only.

### Ethics Statement

Formal ethical approval was not sought for this study as it relied on data that cannot be directly linked to individual subjects.

## Results

### Description of the target population estimate trends

We visualised the trends of the CCPPM, census-based and the 2 EPI numerator as denominator target population estimates from 1999 to 2023 and ANC-1 from 2020-2023 as shown on Figure 1. All the target population methods showed a predominantly upward trend during that period.

**Figure 1:**
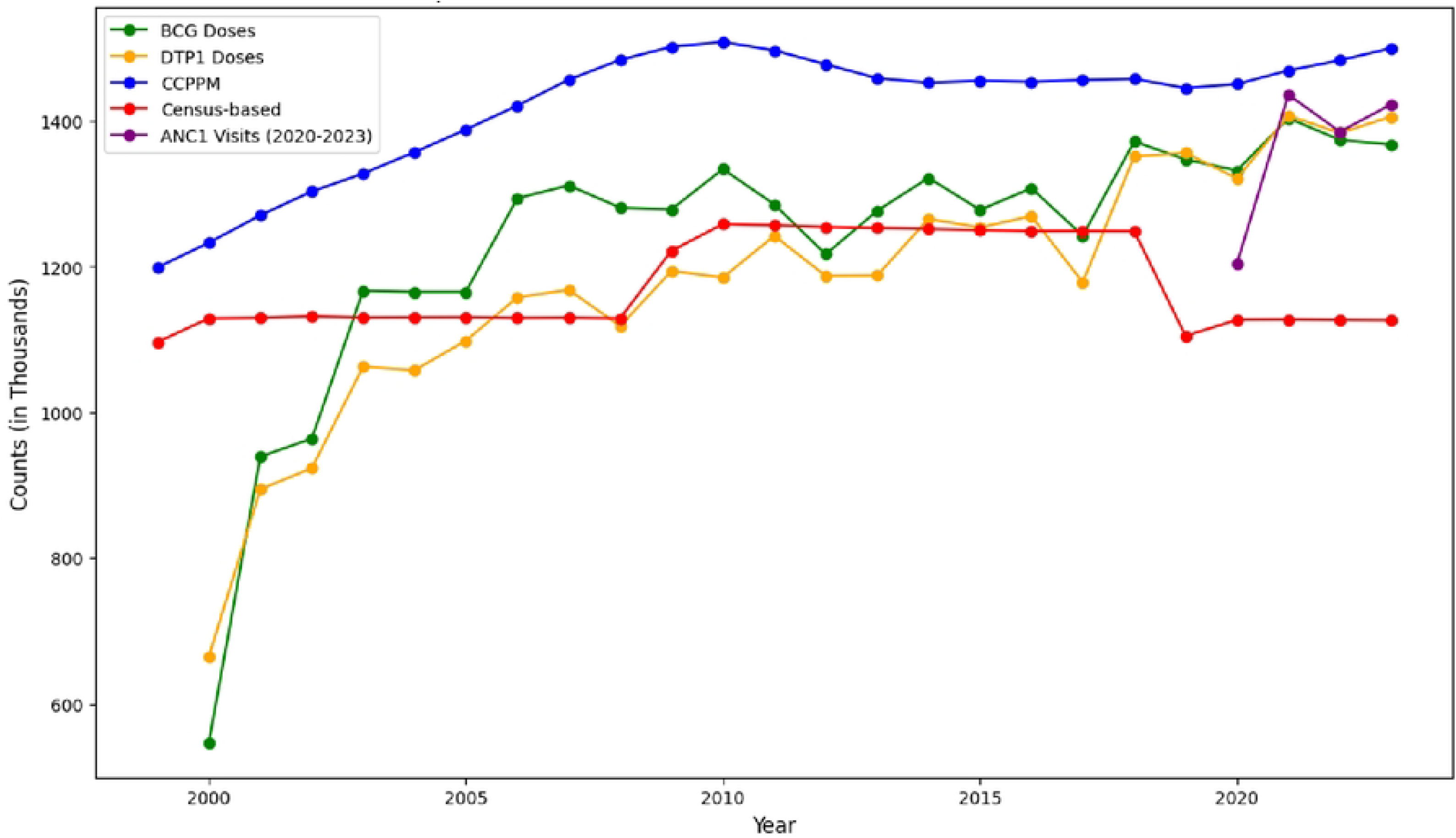
Target population estimates from 1999 to 2023

The CCPPM method had the highest population estimates with a smooth curve throughout the period. In the 2 years following the 1999 census, this estimate was almost similar to the census-based method. During these 2 years, the widest disparity between the CCPPM method and the BCG and the DTP-1 estimates was noted. Following the 2019 census, the EPI numerator as denominator methods were seen to closely match the CCPPM method both in trend and count. In contrast, the census-based estimates showed the greatest divergence in comparison to the CCPPM method after the 2019 census.

The 2 EPI numerator (BCG and DPT-1 doses) as denominator plots showed high congruence in their trends over the 3 census periods. They were at their lowest points in the year 2000, rising steeply over the next 3 years with the BCG curve showing a higher estimate for most of the period, and DTP-1 catching up from 2018 and surpassing BCG in 2023. The plots are not smooth because the estimates are not projections but actual counts of the number of vaccine doses.

The census-based population estimates had 2 points with trend discontinuity interrupting the fairly linear and smooth curve - a sudden population rise in 2009 and a sharp dip in 2019 which were the national census years.

A predominantly upward trend was associated with the CCPPM, BCG, DTP1 and ANC1 methods. The BCG, DTP1 and ANC1 methods showed an almost matching pattern across the years particularly from 2021. The census-based method for that period diverged significantly from the other variables and showed a flat trend. It was therefore excluded from the subsequent analysis.

### Magnitude of error and Linear correlation associated with target population estimate methods

Table 1 shows the MAE, MAPE and Pearson’s Correlation between pairs of the target population estimate methods we investigated. The Mean Absolute Error (MAE) shows the actual difference in count between 2 populations providing a measure of the magnitude of error. The mean absolute percentage error (MAPE) shows the relative degree of error between the 2 populations.

**Table 1:** Mean Absolute Error (MAE), Mean Absolute Percentage Error (MAPE) and Pearson Correlation Coefficient (r) associated with the target population estimation methods (2003-2018)

| Method A | Method B | MAE | MAPE (%) | R |
| --- | --- | --- | --- | --- |
| UNDP | Census-Based | 193756.22 | 16.21 | 0.69 |
| BCG | Census-Based | 69500.82 | 5.39 | 0.49 |
| DTP1 | Census-Based | 44317.16 | 3.77 | 0.76 |
| BCG | DTP1 | 82425.12 | 6.52 | 0.80 |
| CCPPM | BCG | 129772.06 | 10.33 | 0.72 |
| CCPPM | DTP1 | 212917.19 | 18.18 | 0.65 |
| CCPPM | ANC1 | 77574.25 | 5.45 | 0.76 |
| BCG | ANC1 | 56318.25 | 4.16 | 0.89 |
| DTP1 | ANC1 | 40870.25 | 3.04 | 1.00 |

The highest magnitude of error (MAE = 212917.19 and MAPE = 18.18) was observed between the UNDP and DTP1 methods while the lowest magnitude of error was seen between the DTP1 and census-based methods which had MAE = 44317.16 and MAPE = 3.77. The 2 EPI numerator as denominator methods in addition had the highest Pearson Correlation Coefficient of 0.80 which being close to 1, shows a high linear correlation.

The BCG doses compared to the census-based method had a moderate linear correlation (r = 0.49) despite having the second lowest magnitude of error (MAE = 6955.82 and MAPE = 5.39).

The CCPPM compared to the ANC1 method showed the highest magnitude of error (MAE = 77574.25 and MAPE = 5.45) while the lowest was associated with the BCG doses compared to DTP1 doses (MAE = 15448.00 and MAPE = 1.13). A moderate degree of error (MAE = 40870.25 and MAPE = 3.04) was observed between the DTP1 doses and ANC1 methods which had the highest linear correlation (r = 1.00). There was no linear relationship between the CCPPM and BCG methods (r = 0.38). The other paired methods had moderate linear correlation.

## Discussion

Obtaining accurate target population estimates for routine immunisation coverage calculation is challenging in low-income countries including Kenya. Records of births in civil registries are the most accurate source of target population data [14]. Unfortunately, birth registration rates in Kenya are sub-optimal, necessitating the use of census extrapolations to estimate the target population. Census based birth estimates which form the denominator for immunisation coverage calculations tend to be under-estimates, at times yielding coverages of >100%.

In this study, we sought to demonstrate the degree of error associated with the census-based population estimation method compared to the CCPPM, the EPI numerator as denominator and the ANC1 methods. We also aimed to identify an alternative data source that can be considered as a more accurate method of obtaining target population estimates The CCPPM estimates were consistently higher than both the census based and the EPI numerator as denominator methods. This is similar to the findings in 2016 by Kaiser et al, when they compared the trends of reported births and surviving births up to 1 year of 19 eastern and southern African countries from 2000-2013. The pooled data showed that the JRF-reported populations tended to be lower than the CCPPM projections [14]. This has implications on calculated immunisation coverages that then tend to be erroneously higher due to the falsely low denominator.

Following the most recent national census (2019), the census-based target population estimates showed increased divergence from the other methods, with the estimate being even lower than the previous years. Attendance of the first Antenatal care clinic (ANC1) to the analysis showed a similar upward trend to the EPI numerator as denominator and CCPPM methods. This latter finding is surprising and differs from that demonstrated in previous studies that showed a reduction in percentage difference in population estimates over time, with the census -based population estimates becoming more similar to the CCPPM projections. Stashko et al in 2019 compared the target population trends of the 194 WHO member states as reported in the WHO/UNICEF JRF database from 2000-2016 with CCPPM estimates. They noted that population differences became less over time, pointing to a possible improvement in accuracy of target population estimation by countries [15].

The sub-analysis of target population trends from 2020-2023 showed a linear correlation and fairly low magnitude of error between the ANC1 and DTP1 doses. This encouraging observation may imply that health service delivery has improved over time with more women accessing antenatal care services and subsequently taking their children for vaccination. Galadima et al in 2021 reported that women who attend antenatal clinics were more likely to get their children vaccinated [15]. This finding may also be as a result of the improvement in the quality of healthcare services data reporting over time.

The drop in the number of BCG compared to DTP1 vaccine doses in 2022-2023 was most likely due to the prolonged BCG vaccine stock-out during that period [16]. This finding further underscores the need to use 2 closely matched health indicators when estimating a target population for a health intervention so as to mitigate the risk of inaccuracies associated by reliance on only one data set.

This study demonstrated that in the absence of complete birth registration data, it is possible to improve the accuracy of target population estimates using other methods that assess the same population. Utilisation of these alternative methods will serve to improve coverage reporting for immunisation allowing programs to allocate resources efficiently and make timely interventions when coverage drops are observed.

The primary limitation we encountered in this study was missing data. This affected analysis necessitating removal of outliers in one instance and limiting the period where we included ANC1 data to 4 years. Producing high quality health indicator data as well as digitising it on a platform easily accessible to researchers and other key stakeholders is crucial for generating reliable evidence to inform health policy decisions. Another limitation was the fact that we confined our study to Kenya. The findings are therefore not generalizable to other countries as each country has its own unique and context specific demographic and health variables.

One key question that this study did not address was the challenge in obtaining accurate target populations for immunisation faced at sub-national levels. Mapping the performance of individual counties would be important in order in narrowing down the interventions and limited resources on the struggling counties. We would also recommend that mid-census household surveys be conducted every 5 years so as to be able to make adjustments to population estimates in the intercensal period, ensuring more accuracy in the denominator for calculating immunisation coverage.

## Data Availability

The data used in the study is available in the manuscript and figure uploaded. The sources are cited appropriately

